# A structured approach to evaluating life course hypotheses: Moving beyond analyses of exposed versus unexposed in the omics context

**DOI:** 10.1101/19007062

**Authors:** Yiwen Zhu, Andrew J. Simpkin, Matthew J. Suderman, Alexandre A. Lussier, Esther Walton, Erin C. Dunn, Andrew D.A.C. Smith

## Abstract

**Background:** Life course epidemiology provides a framework for studying the effects of time-varying exposures on health outcomes. The structured life course modeling approach (SLCMA) is a theory-driven analytic method that empirically compares multiple prespecified life course hypotheses characterizing time-dependent exposure-outcome relationships to determine which theory best fits the observed data. However, the statistical properties of inference methods used with the SLCMA have not been investigated with high-dimensional omics outcomes.

**Methods:** We performed simulations and empirical analyses to evaluate the performance of the SLCMA when applied to genome-wide DNA methylation (DNAm). In the simulations, we compared five statistical inference tests used by SLCMA (n=700). For each, we assessed the familywise error rate (FWER), statistical power, and confidence interval coverage to determine whether inference based on these tests was valid in the presence of substantial multiple testing and small effect sizes, two hallmark challenges of inference from omics data. In the empirical analyses, we applied the SLCMA to evaluate the time-dependent relationship of childhood abuse with genome-wide DNAm (n=703).

**Results:** In the simulations, selective inference and max-|*t*|-test performed best: both controlled FWER and yielded moderate statistical power. Empirical analyses using SLCMA revealed time-dependent effects of childhood abuse on DNA methylation.

**Conclusions:** Our findings show that SLCMA, applied and interpreted appropriately, can be used in the omics setting to examine time-dependent effects underlying exposure-outcome elationships over the life course. We provide recommendations for applying the SLCMA in high-throughput settings, which we hope will encourage researchers to move beyond analyses of exposed versus unexposed.

**Key messages:**

1. The structured life course modeling approach (SLCMA) is an effective approach to directly compare life course theories and can be scaled-up in the omics context to examine nuanced relationships between environmental exposures over the life course and biological processes.
2. Of the five statistical inference tests assessed in simulations, we recommend the selective inference method and max-|*t*|-test for post-selection inference in omics applications of the SLCMA.
3. In an empirical example, we revealed time-dependent effects of childhood abuse on DNA methylation using the SLCMA, with improvement in statistical power when accounting for covariates by applying the Frisch-Waugh-Lovell (FWL) theorem.
4. Researchers should assess p-values in parallel with effects sizes and confidence intervals, as triangulating multiple forms of statistical evidence can strengthen inferences and point to new directions for replication.

## Introduction

Epidemiologists have long been interested in whether and how exposures over the life course affect later health outcomes. Guided by theories developed in life course epidemiology^1^ (**Table 1**), researchers are moving beyond simple comparisons of the presence versus absence of exposure to characterize time-dependent exposure-outcome relationships.^2^ Prior work in life course epidemiology has conceptualized timing effects in numerous ways, examining the role of the developmental timing of exposure (sensitive period hypothesis), the number of occasions exposed across time (accumulation of risk hypothesis), proximity in time to exposure (recency hypothesis), and change in exposure status across time (mobility hypothesis). Researchers have adopted this life course perspective, uncovering mechanistic insights that advanced many subfields of public health and medicine.^7–11^ As different life course hypotheses correspond to distinct theories of disease etiology, efforts to formally compare competing hypotheses and identify those best supported by empirical data are needed to guide prevention and intervention planning.

**Table 1.**
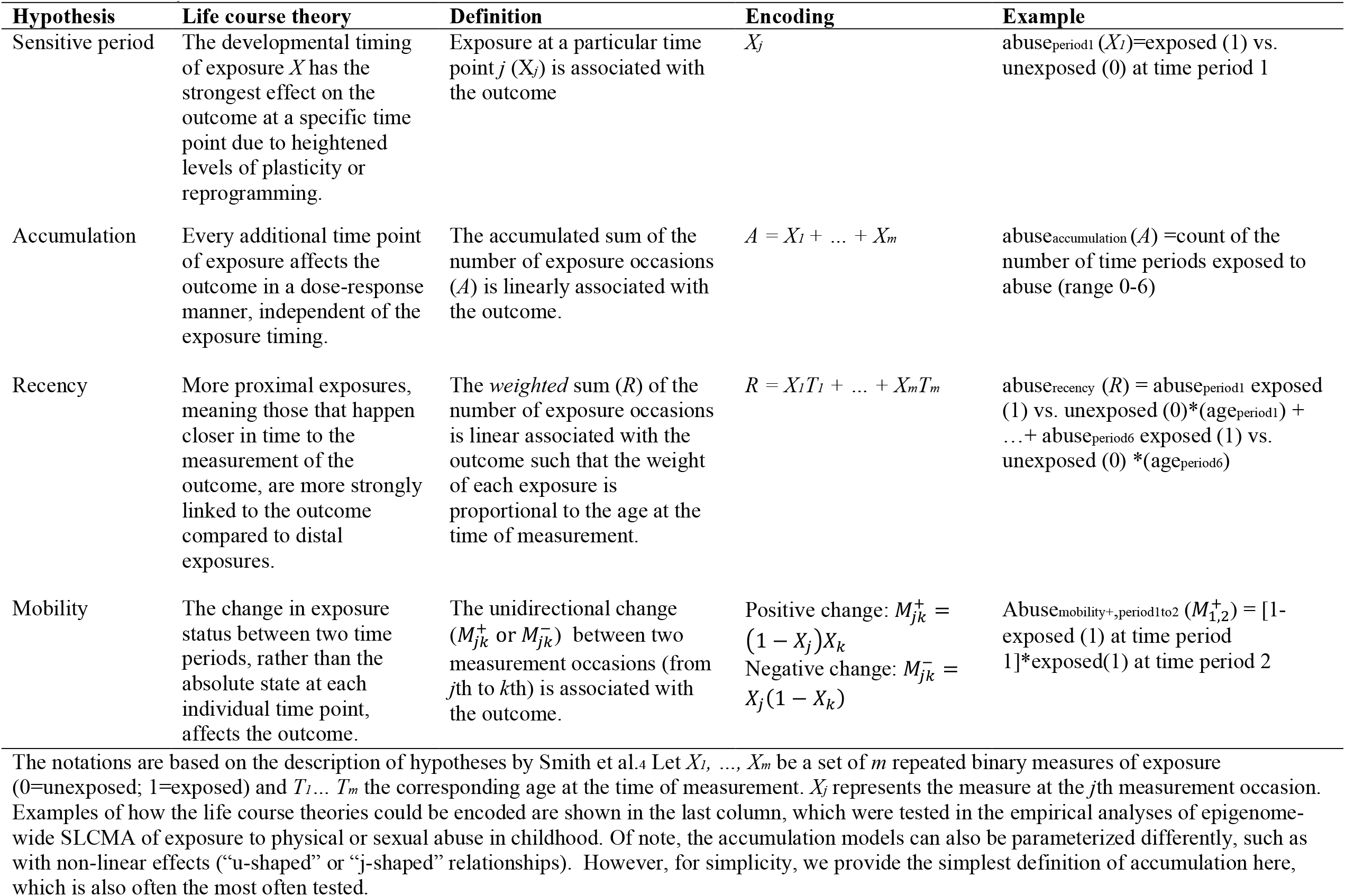
Commonly tested life course theories.

To address the need for systematic comparisons of life course theories, Mishra and colleagues introduced the structured life course modeling approach (SLCMA).^3^ The SLCMA allows researchers to compare a set of a priori-specified life course theories and use goodness-of-fit criteria to determine which theory is best supported by the empirical data. Smith and colleagues later extended this approach by outlining an alternative statistical model selection strategy that makes use of least angle regression (LARS),^12^ accommodates both binary and continuous exposures, ^4,5^ and improves the accuracy of selecting the correct hypothesis. More recently, Madathil et al. proposed a Bayesian approach to life course modeling that does not perform variable selection, but rather estimates the posterior probability corresponding to each theoretical hypothesis while assessing the relative importance of a series of life course theories altogether. ^13^ Since its inception, the SLCMA has been applied in a wide range of non-omic epidemiologic studies, including those examining the time-dependent impacts of childhood trauma, physical activity, or socioeconomic position on psychological, metabolic, and disease outcomes.^6,14–19^

The growing availability of high-dimensional biological and phenotype data from longitudinal cohort studies has created new opportunities to test life course theories in epigenomics, transcriptomics, metabolomics, and other omics settings. While large cross-sectional omics studies have identified *associations* between biological differences and various traits,^20^ applications of the SLCMA to longitudinal data and high dimensional outcomes allow researchers to answer more complex questions about disease *mechanisms*, including: Are there sensitive periods when environmental exposures induce more underlying molecular changes compared to those occurred during other time periods? Do exposures form a dose-response relationship, regardless of their timing? Do more proximal events matter more compared to distal events? As an example, Dunn and colleagues applied the SLCMA in a longitudinal birth cohort study to model timing effects of childhood adversity on DNA methylation, which is a widely studied epigenetic mechanism that could give rise to altered gene expression and phenotypic changes. Using the SLCMA, they found that DNAm differences were largely explained by the age at exposure, with the first three years of life appearing to be a sensitive period associated with more DNAm differences. Interestingly, the SLCMA found associations not uncovered in EWAS of exposed versus unexposed to childhood adversity.^22^

As outlined in Dunn et al.,^22^ application of the SLCMA to omics data presents unique challenges that have not been systematically investigated. First, it remains unknown whether theoretical properties of statistical inference, such as Type I error (i.e., family-wise error rate (FWER) in the presence of multiple testing) or confidence interval (CI) coverage, are valid in omics data. Second, it is unclear whether the SLCMA is sufficiently powered to detect the small effects commonly found in omics settings. Third, questions exist on how to balance decision-making regarding research evidence; omics studies often rely on p-value thresholds whereas epidemiologists and others increasingly prioritize other statistical evidence, such as effect sizes and CIs.^23,24^ We therefore performed both simulations and empirical analyses to assess the performance of the SLCMA when applied to omics data and illustrate how SLCMA can be applied to evaluate the time-dependent role of childhood abuse on genome-wide DNA methylation.

## Methods

### The Structured Life Course Modeling Approach (SLCMA): An Overview

The SLCMA has been described in detail elsewhere.^3–5^ In brief, the SLCMA is a two-stage method that compares a set of life course hypotheses describing the relationship between exposures assessed over time and some outcome of interest. In the first stage of the SLCMA, each life course hypothesis is encoded into a predictor or set of predictor variables. Examples of the predictors that represent commonly studied life course hypotheses are shown in **Table 1**. A variable selection procedure is then used to select the subset of predictors that explain the greatest proportion of outcome variation. While it is possible for multiple predictors to be selected, the high dimensionality of the omics setting makes consideration more feasible of simple life course hypotheses (meaning those in which the exposure-outcome association is represented by a single predictor). Therefore, in this study, we focused on statistical inference regarding the single predictor that explains the greatest variation in the outcome.

In the second stage of the SLCMA, post-selection inference is performed to obtain point estimates (of the direction and magnitude of effects) and confidence intervals (a measure of effect uncertainty) of the effects for the hypotheses identified from the first stage. Post-selection inference methods are used to derive unbiased test statistics because they account for the multiple testing that occurs when comparing multiple hypotheses, as the SLCMA iteratively works to *select* the variable with the strongest association with the outcome. Four primary inference methods that account for this “selective nature” are: (1) Bonferroni correction; (2) max-|*t*|-test;^25^ (3) covariance test;^26,27^ and (4) selective inference.^28,29^ These approaches are described in detail in the **Supplementary Materials.**

### Simulation Analyses

We performed simulations to examine the performance of these four post-selection inference methods compared to a naïve calculation (summarized in **Table 2**). To build these simulations in the context of a real-world application, we modeled the simulation strategy based on the genome-wide SLCMA study performed by Dunn et al.^22^ We evaluated each post-selection inference method with respect to three statistical properties: family-wise error rate (FWER) (the probability of making one or more false discoveries out of multiple tests), statistical power (the probability of correctly selecting the predictor with a true association with the outcome), and CI coverage (the probability that a 95% CI contained the true effect estimate). Assessing these properties enabled us to determine whether inference based on these tests was valid in the presence of multiple testing and small effect sizes, which are two hallmarks of high-dimensional data. Mathematical definitions of the test-statistics as well as the procedure for constructing confidence intervals are included in the **Supplemental Materials**. Example R code for implementing all methods described in this study are also provided in the **Supplemental Materials**.

**Table 2.** Summary of the simulations study setup.

| Under the null (Family wise error rate) |  |  |  |
| --- | --- | --- | --- |
|  | Predictors | Outcome | Number of tests |
| Normal outcomes | Based on exposure to childhood abuse from ALSPAC.<br>Seven variables encoding sensitive period,<br>accumulation and recency hypotheses. | $y \sim \mathcal{N}(0,1)$ | 485 000 |
| Empirical outcomes |  | Resampled DNAm values | 485 000 |
| Under the alternative (Power and confidence interval coverage) |  |  |  |
|  | Predictors | Outcome | Effect size |
| Normal outcomes | Based on exposure to childhood abuse from ALSPAC.<br>Seven variables encoding sensitive period,<br>accumulation and recency hypotheses. | Simulated normal variables associated with the<br>first predictor (earliest sensitive period) | R<sup>2</sup>: 0.01 to 0.1 |
| Empirical outcomes |  | Simulated beta variables associated with the<br>first predictor (earliest sensitive period) | Δ<sub>DNAm</sub>: 0.05 to 0.5 |

The table is divided into two approaches: to assess the familywise error rate, we simulated the exposures and outcomes to have no association with each other (i.e., under the null hypothesis), and ran a single simulation of 485 000 tests to examine the distributions of observed p-values against the expected distribution. To assess the power and confidence interval coverage under the alternative hypothesis, we ran 2 000 simulation experiments to allow the confidence interval (CI) of the assessed metrics (i.e., power and CI coverage) to have a radius (i.e., margin of error) of 1%, setting $\alpha$ to 5%. The two metrics of effect sizes were different with normal versus empirical outcomes due to the difference in the underlying data generating processes.
Sample size was set to $n=700$ in all simulations based on the sample size of the empirical study.
$R^2$ : variance of the outcome explained by the selected predictor.
$\Delta_{\text{DNAm}}$ : difference in average methylation levels between the exposed and unexposed individuals.

We considered two scenarios, which differed in the nature of the simulated outcome:

#### Scenario 1: Normal outcomes

In the first scenario, we simulated exposure to childhood sexual or physical abuse based on empirical data from the Avon Longitudinal Study of Parents and Children (ALSPAC), a population-based birth cohort.^30–32^ The outcome was simulated from a normal distribution and the sample size was set to 700 to be consistent with ALSPAC. Simulations were based on *m*=485 000 tests corresponding to an analysis of Illumina Methylation 450k Beadchip data and a standard genome-wide significance threshold of p<10^−7^. To assess FWER under the null hypothesis, we ran a single simulation of 485 000 tests and examined the distributions of observed p-values against their expected distribution. To assess statistical power and CI coverage, we ran simulations in which the outcome variable was correlated with one of the predictors and then varied the correlation between outcome and predictor such that the variance explained by the predictor varied from 0.01 to 0.1. This range of values was chosen to illustrate poor and good statistical power, respectively.

#### Scenario 2: Empirical outcomes

To further examine the performance of post-selection inference methods in an empirical setting, we combined real predictors from ALSPAC capturing repeatedly measured exposure to childhood sexual or physical abuse as described above and an outcome variable that more closely resembled the data distribution of DNAm at a single locus (meaning the proportion of cells in which the cytosine at the locus is methylated). To assess the FWER under the null hypothesis, we resampled the real predictors and DNAm values from ALSPAC separately. The resampling breaks the predictor-outcome link and hence removes any observed association between the two, while maintaining the empirical distributions of DNAm, which may not be normal. To assess statistical power and CI coverage, we simulated the outcome from a beta distribution, as proposed by Tsai and Bell.^33^ Effect sizes were parameterized as the difference in mean levels of DNAm between the exposed and unexposed groups (*Δ*_*DNAm*_), ranging from 0.05 to 0.5. The number of tests and p-value threshold were the same as Scenario 1. We additionally considered a transformation of the DNAm values from beta values (*y*) to *M* values equivalent to *M*=log_2_(*y*/(1−*y*)), which are sometimes used to stabilize variance.^34^

### Empirical Analyses

To illustrate how the SLCMA and the different corresponding post-selection inference methods work in practice, we reanalyzed data used by Dunn et al.^22^ Briefly, we compared the effects of sensitive period, accumulation, and recency hypotheses for the associations between exposure to sexual or physical abuse and genome-wide DNA methylation at age 7 in ALSPAC participants. Sample characteristics and adversity measures are described in **Supplemental Materials**. Building from that study, which only used the covariance test, we additionally applied the other three post-selection inference methods summarized earlier. We also tested a new method for covariate adjustment that could be used alongside any post-selection inference method. Based on the Frisch-Waugh-Lovell (FWL) theorem, this method regresses the outcome on covariates in parallel and enters the residuals into the model selection procedure.^35–37^ A more thorough description of this method and a full list of the covariates are available in the **Supplemental Materials**.

## Results

### Simulation Analyses

**Table 3** summarizes the main findings from the simulation analyses with regard to the statistical properties and implementation of the assessed methods.

#### Family-wise error rate

Due to the high computational burden of genome-wide association studies, we illustrate FWER control of each inference test using a single simulation with *m*=485 000 tests. As shown in **Figures 1 and 2**, when compared against the expected p-value distribution under the null hypothesis, the p-values obtained from naïve calculations appeared too liberal in both scenarios, as suggested by the systematic upward departure from the diagonal line. P-values from the covariance test were also smaller than expected across scenarios.

**Table 3.** Summary of main findings: statistical properties of post-selection inference methods.

| **Method** | **FWER**<br>**(Figures 1 & 2)** | **Statistical power**<br>**(Figure 3)** | **CI coverage**<br>**(Figure 4)** | **Software**<br>**availability** | **Computation time**<br>**for an epigenome-**<br>**wide analysis\*** |
| --- | --- | --- | --- | --- | --- |
| Naïve calculation | Inflated p-values<br>and FWER | Biased due to<br>inflated FWER | Lower than<br>expected coverage<br>when effect size is<br>small<sup>4</sup> | Widely available | Fast (24 minutes) |
| Bonferroni correction | Controlled at any<br>level | Comparable | Overconservative<br>(i.e., above<br>expected coverage) | Widely available | Fast (24 minutes) |
| Max-$\vert t\vert $-test | Controlled at any<br>level | Comparable | Lower than<br>expected coverage<br>when effect size is<br>small | R code provided in<br>the Supplement | Slow (11 hours 51<br>minutes) |
| Covariance test | Inflated p-values<br>and FWER | Biased due to<br>inflated FWER | Expected<br>coverage;<sup>4</sup> interval<br>not necessarily<br>contiguous | R Package<br>archived<sup>27</sup> | Moderate (1 hour<br>19 minutes) |
| Selective inference | Controlled at any<br>level | Comparable | Expected coverage | R Package<br>available;<sup>29</sup><br>possible to<br>implement<br>generalized linear<br>models as well | Slow (14 hours 13<br>minutes) |

FWER: family-wise error rate; CI: confidence interval
\*Computation time was based on analyses running under R 3.4.0 using a high performance computer cluster with 8GB RAM and a maximum of 6 CPU cores allotted.

**Figure 1.**
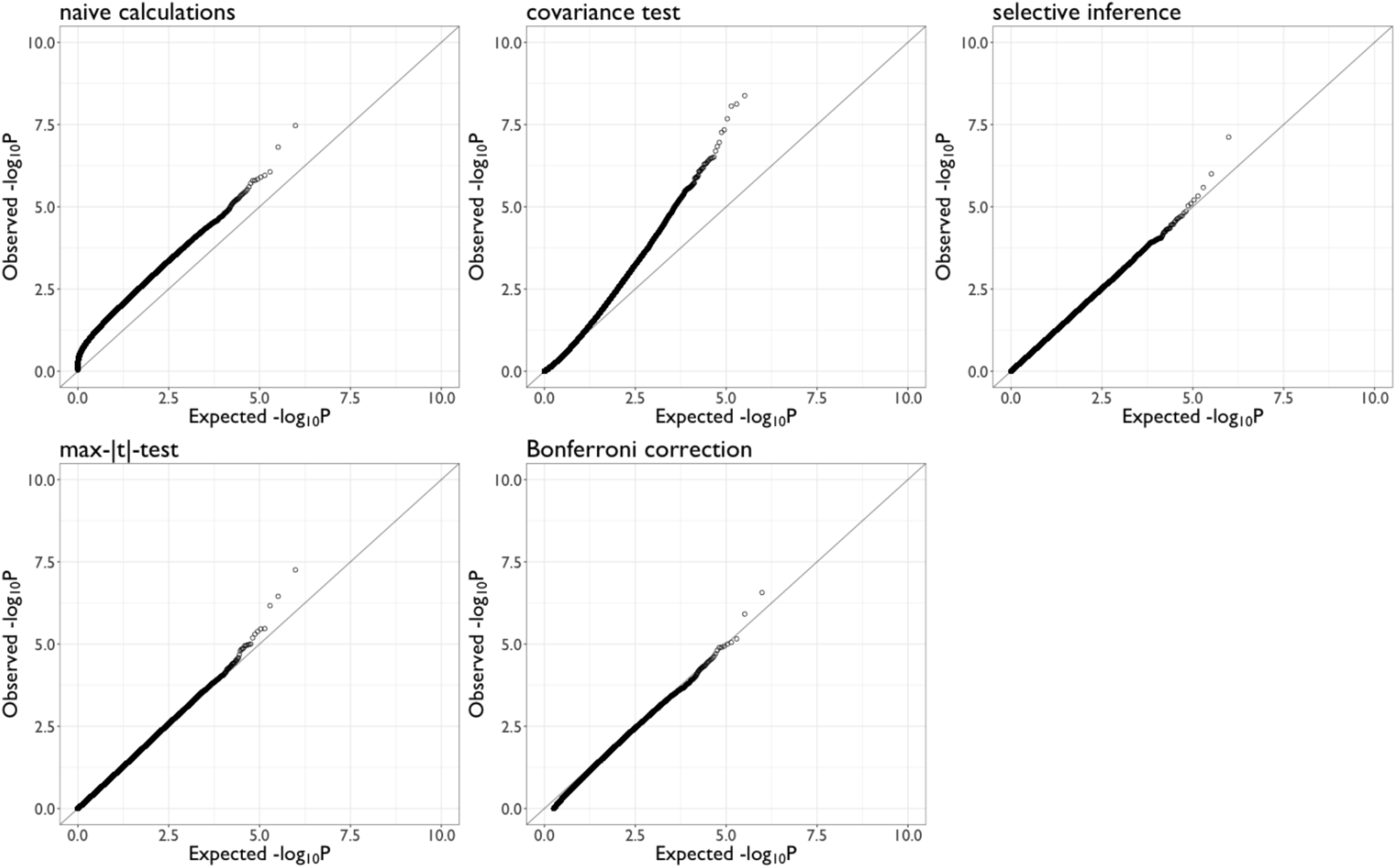
Q-Q plots comparing the expected versus observed p-values simulated under the null for naïve calculations and four post-selection inference methods (N=700) with normal outcomes, where the outcome variables were simulated to follow a normal distribution (scenario 1).

**Figure 2.**
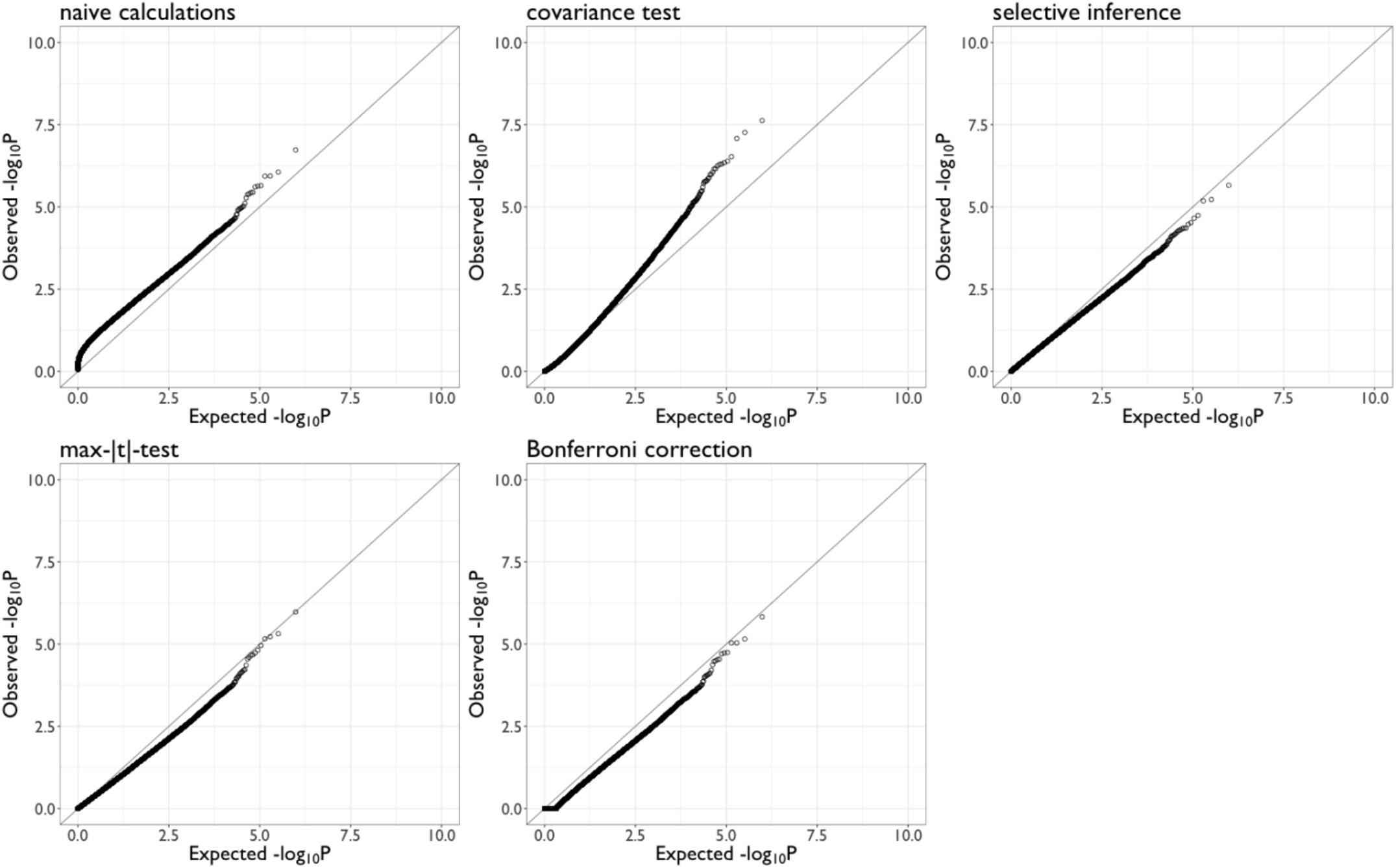
Q-Q plots comparing the expected versus observed p-values simulated under the null for naïve calculations and four post-selection inference methods (N=700) with empirical outcomes, where the outcome variables were resampled from observed DNAm values (scenario 2).

With normally distributed outcomes in Scenario 1, the p-values from the Bonferroni correction, max-|*t*|-test, and selective inference method followed the expected distribution closely (**Figure 1**). With empirical DNAm outcomes in Scenario 2, p-values from the three methods seemed too conservative (**Figure 2**). Transforming the DNAm (beta) values to *M*-values did not affect the results (**Figure S1**). Together, these findings suggest that three methods adequately controlled the FWER: Bonferroni correction, the max-|*t*|-test, and the selective inference method.

Estimates of FWER from repeated simulation experiments when the number of tests ranged from *m*=1 to 1000 are available in the **Supplemental Materials**.

#### Statistical Power and CI Coverage

We assessed the statistical power of the three methods that adequately controlled FWER. We did not evaluate the performance of the covariance test or naïve calculation, as these methods had smaller than expected p-values under the null hypothesis in all simulations. Inclusion of these methods would have thus resulted in an unfair comparison, as their statistical power is inflated by their tendency to fail to reject the null hypothesis.

**Figure 3** shows the estimated statistical power as a function of effect sizes tested both with normal and empirical outcomes. There was very little difference in statistical power between the three methods; they all had ideal statistical power (over 80%) when the effects were moderate to large (*R*^*2*^ > 0.06 in scenario 1; *Δ*_*DNAm*_ *>* 0.25 in scenario 2).

**Figure 3.**
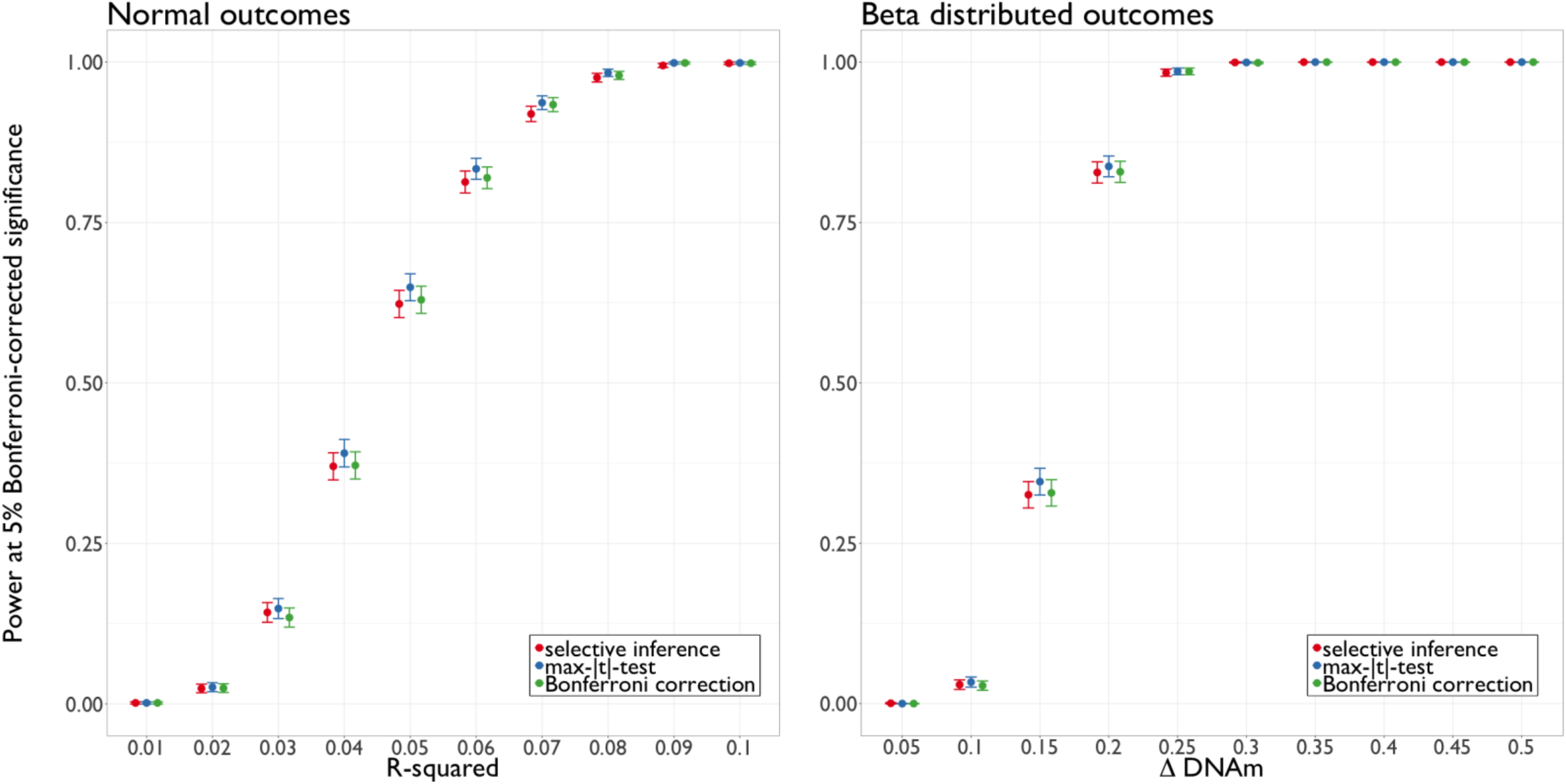
Estimated statistical power and corresponding 95% CI in simulated epigenome-wide analyses (n=700), with varying effect sizes.

**Figure 4** shows CI coverage as a function of effect size. With normal outcomes, the selective inference achieved ideal coverage (around 95%) across all effect sizes with sample size n = 700; the max-|*t*|-test had slightly lower coverage when the effect size was small (*R*^*2*^ < 0.03). With empirical (beta distributed) outcomes, the CI coverage probabilities were below the desired level (95%) when the between-group difference (*Δ*_*DNAm*_) was below 0.3, though exceeded 95% as the effect size increased. Bonferroni corrected CIs were over-conservative across effect sizes and scenarios, as expected.

**Figure 4.**
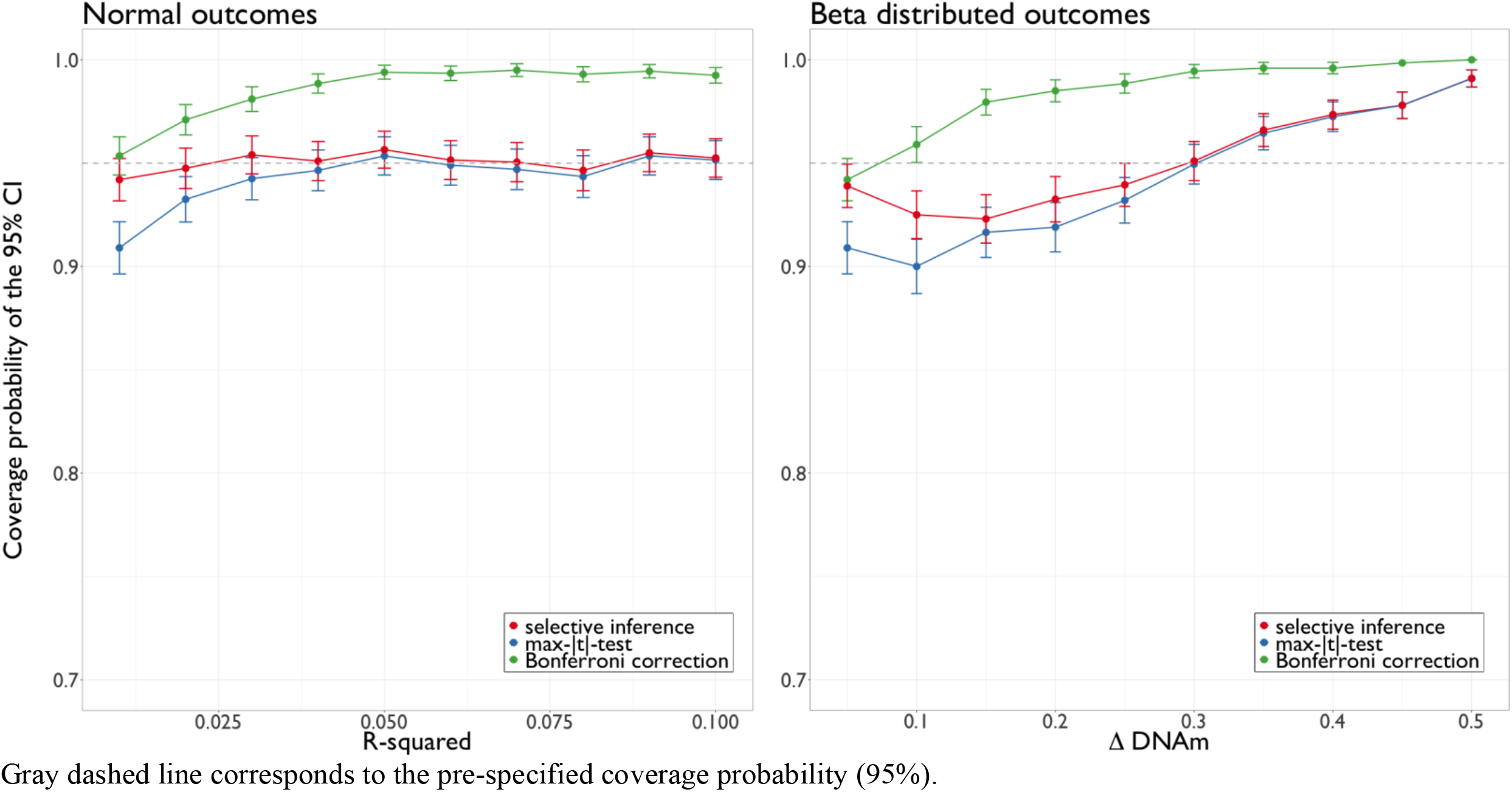
Estimated confidence interval coverage probability and corresponding 95% CI in simulated epigenome-wide analyses (n=700), with varying effect sizes. Gray dashed line corresponds to the pre-specified coverage probability (95%). Gray dashed line corresponds to the pre-specified coverage probability (95%).

### Empirical Analyses

Using the covariance test, Dunn and colleagues identified five CpG sites in ALSPAC that showed differential methylation profiles at age 7 following exposure to physical or sexual abuse in childhood; the “sensitive period” model was the selected life course theory for these five sites. We reanalyzed the genome-wide SLCMA analyses using two other post-selection inference methods that showed no inflation in FWER and desired CI coverage: the max-|*t*|-test and selective inference method. Results are shown in **Table S1**. While neither method identified any CpG site as significantly associated using a stringent Bonferroni corrected p-value threshold of p<1×10^−7^, the CpG site with the smallest p-value from the covariance test (*cg06430102*) remained the CpG with the smallest p-value (out of the 485 000 CpG sites tested) for the two alternative methods (**Table S1**). The CI calculated based on the covariance test, selective inference, and the max-|*t*|-test substantially overlapped (**Figure 5; Table S1**).

**Figure 5.**
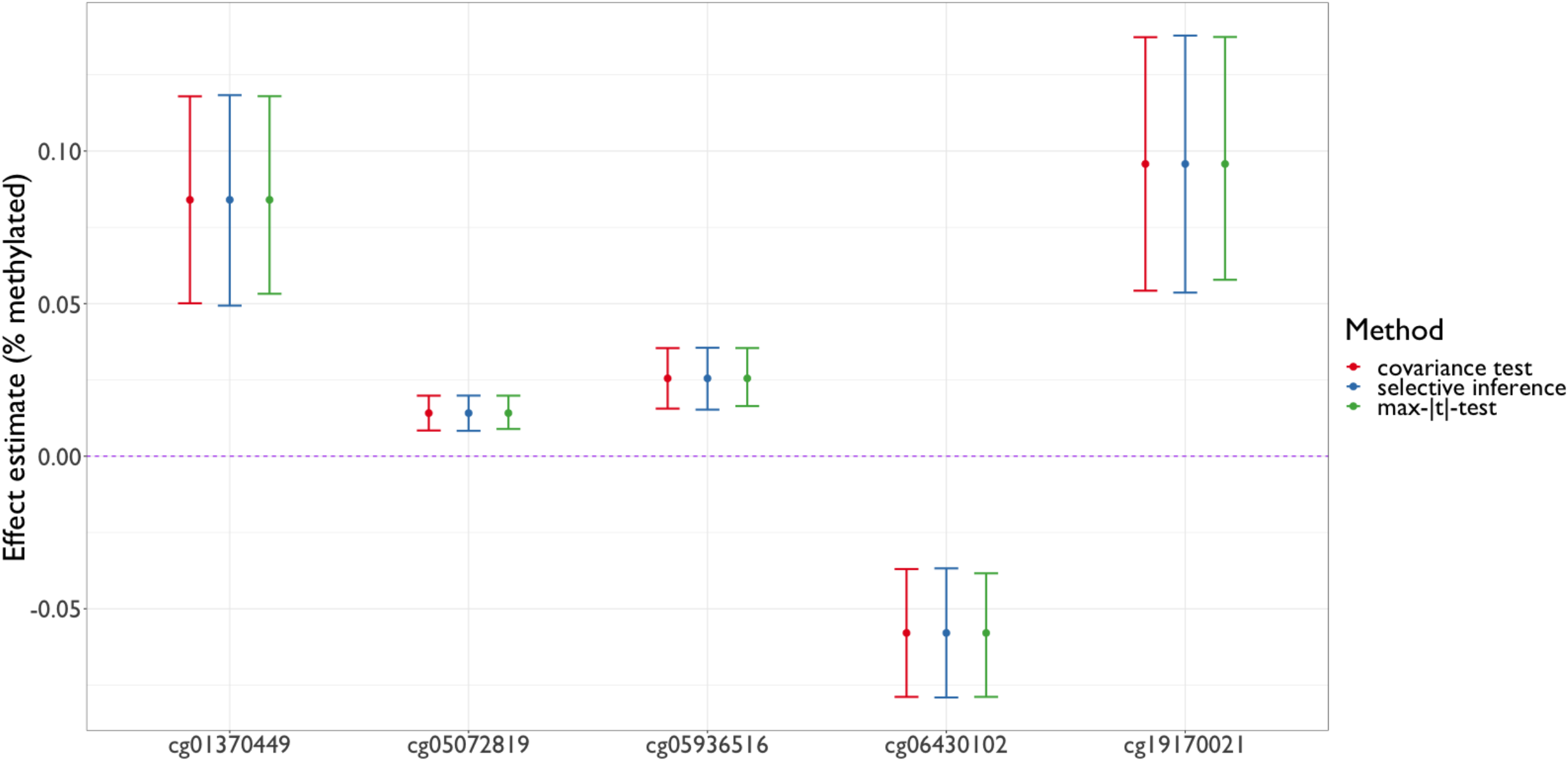
Overlap between confidence intervals based on the covariance test, selective inference, and the max-|t|-test in the empirical example, showing the top five loci.

After applying the Frisch-Waugh-Lovell (FWL) theorem to additionally adjust for covariates, the p-values decreased at all five loci (**Figure S2**), suggesting that the approach improved statistical power and additionally controlled for any bias that may be caused by confounding.

On a genome-wide level, **Table S2** shows the overlap in most strongly associated loci based on results obtained from the three methods assessed in the empirical analyses to compare the preferred alternative methods to the widely-used (but now identified as inflated) covariance test. The concordance between the liberal covariance test and recommended selective inference method was high, implying that both methods agreed on the loci with the strongest associations with the exposure.

## Discussion

As the availability of longitudinal biological and phenotypic data grows, methods from life course epidemiology can be translated to “harness the ‘omics’ revolution” ^7^ and reveal critical insights about how exposures become biologically embedded. We showed that one such method – the SLCMA – is an effective approach to directly compare life course theories and can be scaled-up to answer nuanced questions about complex and time-dependent exposure-omics relationships. Importantly, not all methods for statistical inference in the SLCMA are suitable in high-throughput applications. Based on our findings, we recommend the selective inference method and max-|*t*|-test for post-selection inference in omics applications. Our simulations also showed that statistical power to detect effects depended on effect size, but not necessarily on the post-selection inference method used. When deciding between these two inference methods, researchers will need to consider several factors, including goals of analysis and study-specific contexts, as both methods have strengths and limitations in these areas (**Supplemental Materials**). The simulation analyses highlight the value of using simulations in scientific research,^39,40^ especially when theoretical assumptions may be violated in a new application setting.

In the empirical analyses, we also showed that evaluating multiple forms of evidence can help contextualize the SLCMA findings in omics applications, though statistical significance based on p-values may differ across methods. Researchers should assess p-values in parallel with effects sizes and confidence intervals, as decision rules of significance based on p-values of one method may be biased due to inflation or overcorrection. While it remains challenging to draw definitive conclusions about the presence versus absence of an effect, triangulating evidence from multiple sources and methods may suggest directions for future replication.^41^ For example, a CpG that was identified as the top site by multiple methods and showed substantial changes in methylation levels between exposed versus unexposed individuals may be more likely to capture effects of the exposure and worth pursuing in experimental validation.

Several limitations are noted. First, although we varied the effect size and compared normal versus empirical distributions of the outcome, we did not assess diverse correlation structures or distributions in the exposures tested, as it was not possible given the number of possible combinations of these parameters. Thus, we encourage researchers to perform their own simulations to arrive at a better understanding of the statistical properties of the SLCMA in their specific research context. Second, we restricted our analyses to the application of linear regression-based model selection; a brief discussion on the possibility of implementing post-selection inference methods for generalized linear models is included in the **Supplemental Materials**. Third, as suggested by the simulations, a typical longitudinal epigenetic study with a sample size under 1000 is likely underpowered to detect small effects. One approach to improve statistical power is to combine data or summary results from multiple samples and perform a mega/meta-analysis; developing methods to meta-analyze results from SLCMA analyses is an important goal of future work. Another approach is to use the FWL theorem for covariate adjustment, which as shown in this paper led to improvement in power.

In conclusion, the SLCMA is a useful approach that brings the life course perspective into the omics context. This analysis framework can reveal site-specific mechanisms, as well as system-level pathways. Compared to an analysis that only categorized exposure status as exposed versus unexposed, the SLCMA not only offers additional mechanistic insights, but also increases statistical power when the true underlying exposure-outcome relationship is more nuanced.^22^ As a field, we should move beyond analyses of the presence versus absence of exposure, and make full use of repeatedly measured phenotype and omics data to generate knowledge that improves human health over the life course.

## Data Availability

In the simulations, code used to perform analysis has been included in the Supplemental Materials. In the empirical analysis, data came from the Avon Longitudinal Study of Parents and Children (ALSPAC). More details about data access are available on the ALSPAC website, including a fully searchable data dictionary: http://www.bristol.ac.uk/alspac/researchers/our-
data/.

## Acknowledgements

We are extremely grateful to all the families who took part in the ALSPAC study, the midwives for their help in recruiting them, and the whole ALSPAC team, which includes interviewers, computer and laboratory technicians, clerical workers, research scientists, volunteers, managers, receptionists and nurses. The UK Medical Research Council and the Wellcome Trust (Grant ref: 102215/2/13/2) and the University of Bristol provide core support for ALSPAC. ARIES was funded by the BBSRC (BBI025751/1 and BB/I025263/1). Supplementary funding to generate DNA methylation data which is included in ARIES has been obtained from the MRC, ESRC, NIH and other sources. ARIES is maintained under the auspices of the MRC Integrative Epidemiology Unit at the University of Bristol (MC_UU_12013/2 and MC_UU_12013/8). A comprehensive list of grants funding is available on the ALSPAC website (http://www.bristol.ac.uk/alspac/external/documents/grant-acknowledgements.pdf). This publication is the work of the authors, each of whom serve as guarantors for the contents of this paper.

